# Novel analytical application of the pressure phase plane for evaluating pulmonary artery wave reflection in surgically repaired tetralogy of Fallot

**DOI:** 10.64898/2026.05.09.26352788

**Authors:** Yasunobu Hayabuchi, Yukako Homma

**Affiliations:** Department of Pediatrics, Tokushima University, Tokushima, Japan

**Keywords:** right ventricle, tetralogy of Fallot, pulmonary artery wave reflection, pressure phase plane

## Abstract

**Background:** Pulmonary artery (PA) wave reflection is a key determinant of right ventricular (RV) afterload. RV function is the most important factor determining long-term prognosis in patients with surgically repaired tetralogy of Fallot (rTOF). This study aimed to evaluate PA wave reflection in rTOF using RV pressure phase plane (PPP) analysis, and to identify the clinical, morphological, and hemodynamic characteristics associated with increased PA wave reflection in patients with rTOF.

**Methods:** Augmentation pressure (AugPr) during late systole was quantified using the inflection point of systolic dP/dt on the PPP. The ratio of AugPr to RV systolic pressure (RVSP) was defined as the AugPr index. The study included 87 patients with rTOF (mean age, 15.9 ± 10.0 years), 17 control subjects (13.3 ± 6.3 years), and seven patients with pulmonary arterial hypertension (PAH) (16.4 ± 11.7 years). The rTOF cohort was categorized according to surgical procedure: pulmonary valve–sparing repair (PVS, n = 5), transannular patch repair (TAP, n = 34), and the Rastelli procedure (n = 48).

**Results:** The prevalence of AugPr was 0% in the control group, 100% in the PAH group, and 26.4% in the rTOF group (p < 0.0001). Among the surgical subgroups, the prevalence was 0% in PVS, 14.7% in TAP, and 41.7% in the Rastelli group (p < 0.0027). AugPr and the AugPr index were significantly higher in the Rastelli group than in the other two groups (p = 0.0447 and 0.0433, respectively). In addition, AugPr showed significant correlations with RVSP, RV outflow tract obstruction, maximal dP/dt, and pulmonary regurgitation grade (all p < 0.05).

**Conclusions:** PA wave reflection can be clearly visualized using PPP. The Rastelli group demonstrated a higher prevalence and magnitude of PA wave reflection, suggesting a greater increase in RV afterload compared with other surgical repair types.

## Introduction

Right ventricular (RV) function is recognized as the most important determinant of quality of life and long-term prognosis in patients with surgically repaired tetralogy of Fallot (rTOF) [1–4]. In this population, pulmonary artery (PA) wave reflection is considered a key factor that directly influences prognosis by determining RV afterload [5,6]. PA wave reflection after rTOF is affected by multiple factors, including PA stiffness, RV outflow tract (RVOT) morphology, the presence of prosthetic conduits, branch PA configuration, and patch materials [7–9]. These structural and vascular factors may significantly influence RV afterload. However, to our knowledge, no studies have quantitatively evaluated PA wave reflection in this patient population, and no data have been published clarifying which RV or PA characteristics contribute to the augmentation of PA wave reflection.

In systemic circulation, the magnitude of wave reflection is commonly assessed using the augmentation index (AIx) derived from the aortic pressure waveform, and this parameter has been widely applied in clinical practice [10–12]. The AIx is defined as the ratio of late systolic pressure augmentation (ΔP) to pulse pressure (PP) (ΔP/PP). On the other hand, in patients with rTOF, severe pulmonary regurgitation (PR) is frequently present and substantially affects PA diastolic pressure. Therefore, evaluating wave reflection using PA PP to calculate the AIx is not appropriate in this population. Given these hemodynamic characteristics, we hypothesized that PA wave reflection in patients with rTOF could be more appropriately assessed using the RV pressure waveform. We further considered that pressure phase plane (PPP) analysis of the RV pressure waveform could provide a simpler and more suitable method for this evaluation.

Given this background, the present study aimed to assess the feasibility and clinical utility of quantitative evaluation of PA wave reflection using PPP analysis, and to identify the clinical, morphological, and hemodynamic characteristics associated with increased PA wave reflection in patients with rTOF.

## Methods

### Study population

The study population consisted of 87 patients who had undergone intracardiac rTOF (rTOF group). All patients were scheduled to undergo routine cardiac catheterization for evaluation of their hemodynamic status. According to the surgical technique, the rTOF cohort was classified into three subgroups: pulmonary valve–sparing repair (PVS, n = 5), transannular patch repair (TAP, n = 34), and the Rastelli procedure (n = 48). In the rTOF group, patients were included only if they had no significant residual intracardiac shunt, no severe tricuspid regurgitation, and no dynamic RV outflow tract obstruction (RVOTO). For comparison, seven patients with pulmonary arterial hypertension (PAH group) and 17 control subjects with normal hemodynamics (control group) were also evaluated using the same protocol. The conditions of the patients in the PAH group were as follows: idiopathic PAH (n = 5); idiopathic PAH with a coincidental small atrial septal defect (n = 1); and a small ventricular septal defect (n = 1). The control group consisted of patients with the following diagnoses: nine patients after Kawasaki disease without any coronary arterial stenosis or myocardial ischemia; five patients with patent ductus arteriosus with Qp/Qs < 1.1, for whom catheter occlusion was planned; and three patients who had concealed Wolf–Parkinson–White syndrome and had undergone catheter ablation. Those with left ventricular (LV) and RV pressures, volumes, and function assessed as normal were enrolled in the control group.

Data collected from December 2021 to December 2025 were analyzed. All study protocols conformed to the ethical guidelines of the Declaration of Helsinki (1975) and were approved by the Institutional Review Board of Tokushima University Hospital (approval No.: 1692-3). Written, informed consent for their children to participate in the study was provided by the parents or guardians.

### Cardiac catheterization

Cardiac catheterization and angiography (Integris Allura 9 Biplane; Phillips Medical Systems, Best, The Netherlands) proceeded using 4- to 6-Fr catheters. Data were acquired during routine cardiac catheterization. LV and RV pressure measurements were performed using a high-fidelity manometer-tipped 0.014-inch pressure wire (PressureWire Aeris; St. Jude Medical, St. Paul, MN, USA). Recordings were made with respiration suspended at the end of expiration. All hemodynamic data were acquired at a sampling rate of 100 Hz before the administration of any contrast agents.

### Pressure phase plane (PPP) analysis

In all patients, RV pressure was measured and expressed as time-varying pressure, P(t), during cardiac catheterization (Fig 1A and 1C). The PPP, in which the time derivative of pressure (dP/dt) is plotted against P(t), was then constructed (Fig 1B and 1D) [13–15]. In the PPP representation, one cardiac cycle is depicted as a single clockwise rotation. PPP analysis has previously been shown to be clinically useful for evaluating parameters such as maximal and minimal dP/dt (dP/dt_max and dP/dt_min, respectively), end-diastolic pressure, systolic performance, and diastolic function [13–16].

**Fig 1.**
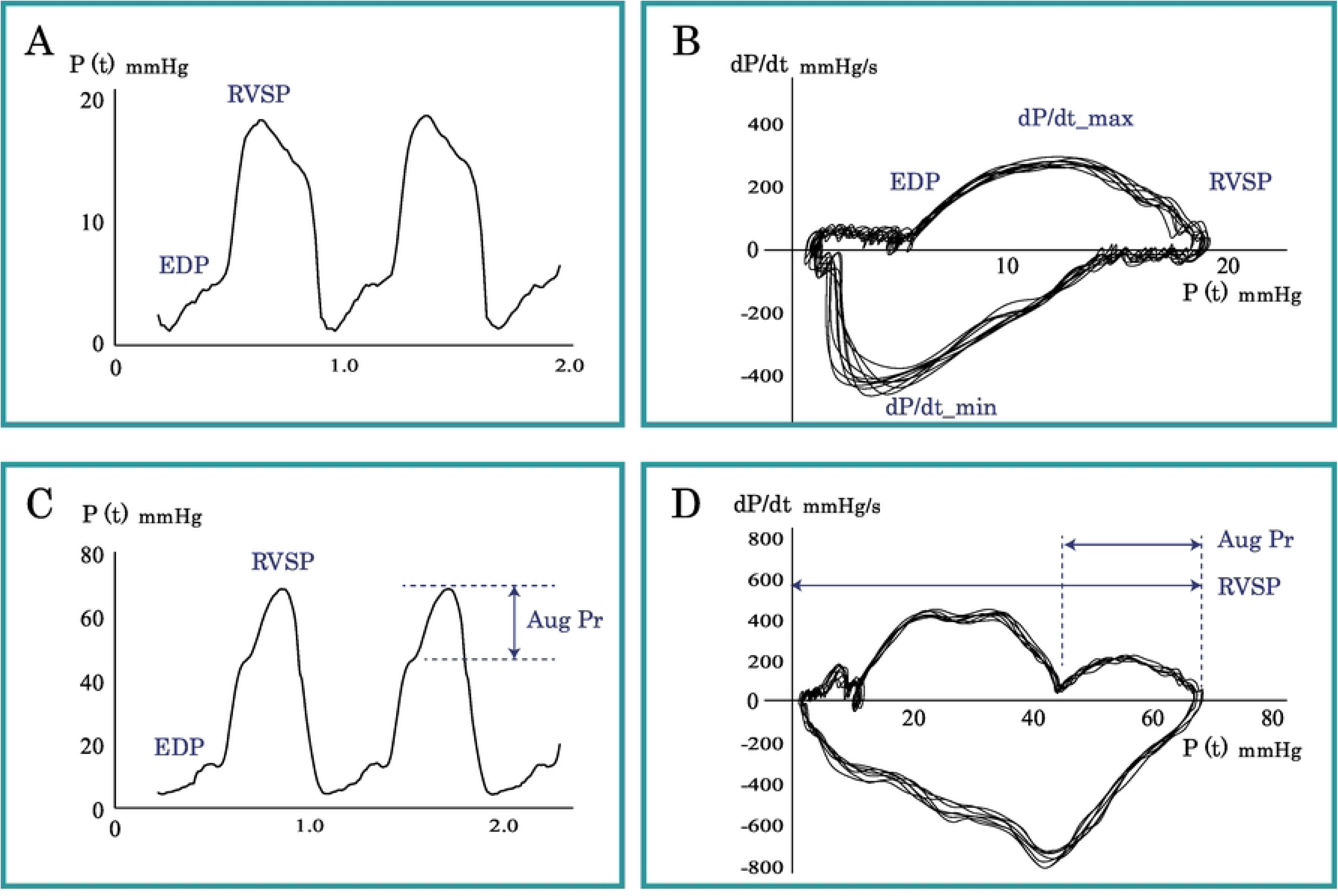
Right ventricular (RV) pressure waveform and Pressure phase plane (PPP) of RV pressure in the control group and he pulmonary arterial hypertension (PAH) group. A. Representative recording of a right ventricular (RV) pressure waveform from a subject in the control group. B. Pressure phase plane (PPP) of RV pressure from the same control subject shown in A. No late systolic pressure augmentation is observed in this case. C. Representative recording of an RV pressure waveform from a patient in the pulmonary arterial hypertension (PAH) group. D. PPP of the RV pressure from the same patient with PAH shown in C. In this case, AugPr is 20 mmHg and the AugPr index is 0.32. RVSP, right ventricular systolic pressure; EDP, end-diastolic pressure; AugPr, augmentation pressure.

### Quantification of pulmonary wave reflection using PPP

The onset of late systolic augmentation pressure (AugPr) was defined as the inflection point of the systolic pressure curve, specifically the point during systole at which dP/dt changed from a decreasing to an increasing trend. In cases in which no such inflection point was identified, late systolic pressure augmentation was considered absent. To assess PA reflection, we defined the AugPr index as follows using RV systolic pressure (RVSP) and AugPr: AugPr index = AugPr / RVSP.

### Statistical analysis

All data are expressed as the mean ± standard deviation (SD) or the median with the 5th to 95th percentiles. The significance of differences was determined using the Mann–Whitney *U* test or the Kruskal–Wallis test followed by Dunn’s test, as appropriate. Linear regression analyses were performed for the correlations, and Pearson’s or Spearman’s correlation coefficients were calculated. All statistical data were analyzed using Prism (version 10.0; GraphPad Software, San Diego, CA, USA) and JMP 19 (SAS Institute, Cary, NC, USA). P-values < 0.05 (two-sided) were considered significant. Intraobserver variability was assessed by one investigator (Ya.H.) conducting measurements on the same patients 8 weeks apart, and interobserver variability was assessed by a second investigator (<u>Yu.H.</u>) who was unaware of the previous results and performed the same measurements on 10 randomly selected participants. Intraobserver and interobserver agreements were assessed using intraclass correlation coefficients (ICCs). In addition, agreement between investigators was tested using Bland–Altman analysis by calculating the bias (mean difference) and 1.96 SD around the mean difference.

## Results

No subjects were excluded from the analysis because of suboptimal pressure recordings. Accordingly, the final study population consisted of 87 patients (mean age ± SD, 15.9 ± 10.0 years) in the rTOF group, seven (mean age ± SD, 16.4 ± 11.7 years) in the PAH group, and 17 (mean age ± SD, 13.3 ± 6.3 years) in the control group. The clinical and hemodynamic characteristics of these three groups are summarized in Table 1.

**Table 1.**
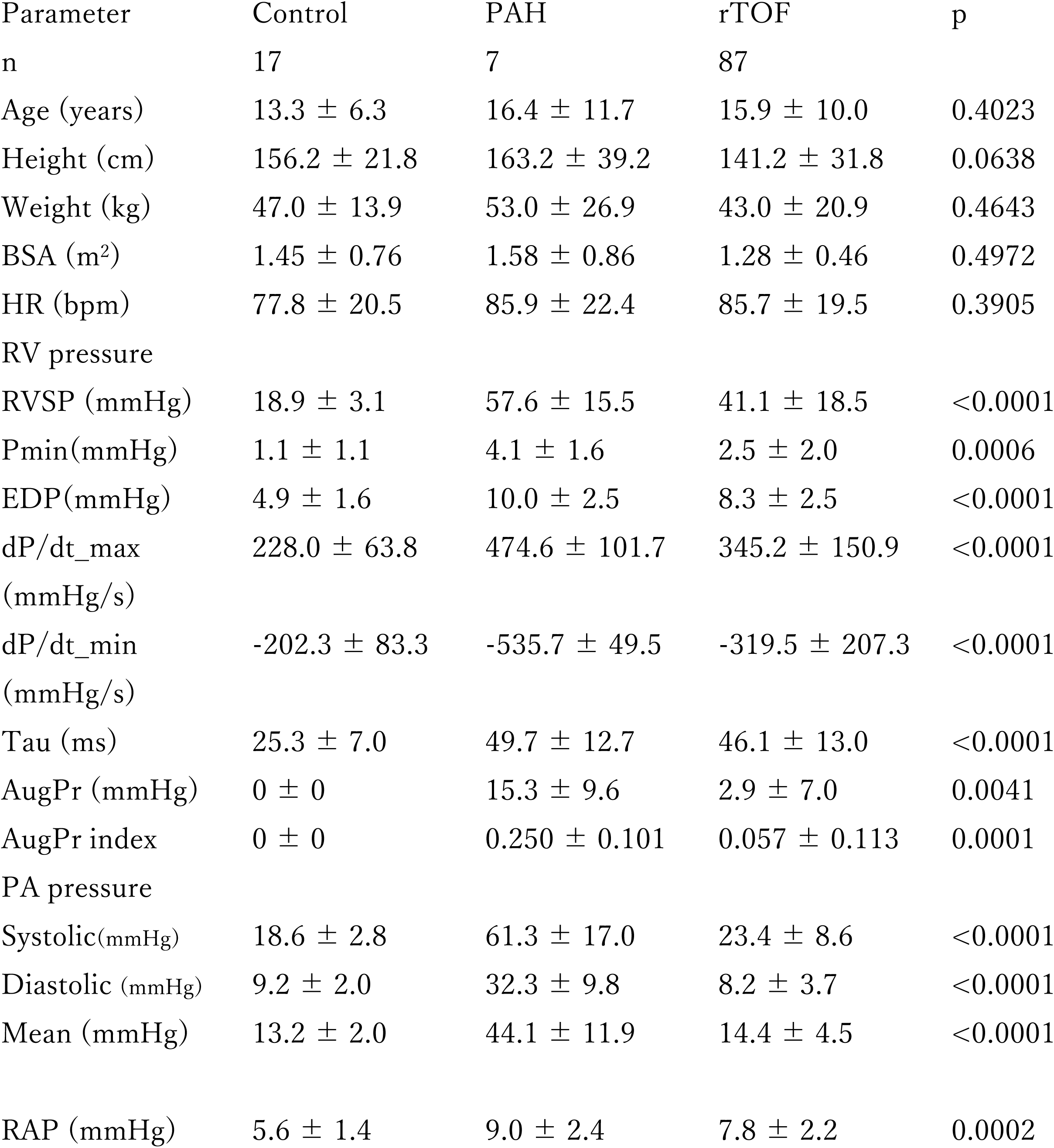
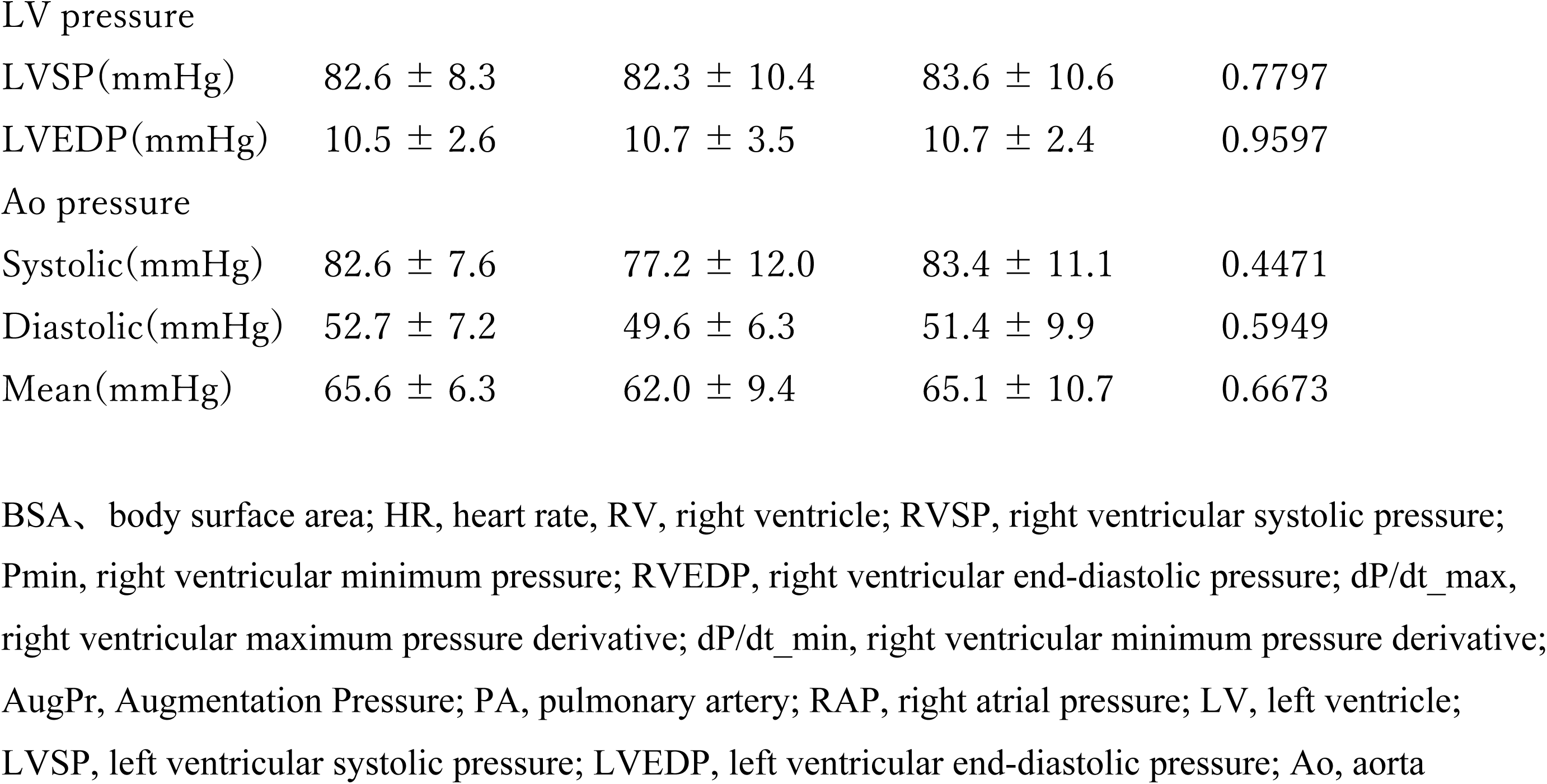
The clinical and hemodynamic characteristics in the Control, PAH, and rTOF groups.

| Parameter | Control | PAH | rTOF | p |
| --- | --- | --- | --- | --- |
| n | 17 | 7 | 87 |  |
| Age (years) | $13.3 \pm 6.3$ | $16.4 \pm 11.7$ | $15.9 \pm 10.0$ | 0.4023 |
| Height (cm) | $156.2 \pm 21.8$ | $163.2 \pm 39.2$ | $141.2 \pm 31.8$ | 0.0638 |
| Weight (kg) | $47.0 \pm 13.9$ | $53.0 \pm 26.9$ | $43.0 \pm 20.9$ | 0.4643 |
| BSA (m<sup>2</sup>) | $1.45 \pm 0.76$ | $1.58 \pm 0.86$ | $1.28 \pm 0.46$ | 0.4972 |
| HR (bpm) | $77.8 \pm 20.5$ | $85.9 \pm 22.4$ | $85.7 \pm 19.5$ | 0.3905 |
| RV pressure |  |  |  |  |
| RVSP (mmHg) | $18.9 \pm 3.1$ | $57.6 \pm 15.5$ | $41.1 \pm 18.5$ | <0.0001 |
| Pmin(mmHg) | $1.1 \pm 1.1$ | $4.1 \pm 1.6$ | $2.5 \pm 2.0$ | 0.0006 |
| EDP(mmHg) | $4.9 \pm 1.6$ | $10.0 \pm 2.5$ | $8.3 \pm 2.5$ | <0.0001 |
| dP/dt_max (mmHg/s) | $228.0 \pm 63.8$ | $474.6 \pm 101.7$ | $345.2 \pm 150.9$ | <0.0001 |
| dP/dt_min (mmHg/s) | $-202.3 \pm 83.3$ | $-535.7 \pm 49.5$ | $-319.5 \pm 207.3$ | <0.0001 |
| Tau (ms) | $25.3 \pm 7.0$ | $49.7 \pm 12.7$ | $46.1 \pm 13.0$ | <0.0001 |
| AugPr (mmHg) | $0 \pm 0$ | $15.3 \pm 9.6$ | $2.9 \pm 7.0$ | 0.0041 |
| AugPr index | $0 \pm 0$ | $0.250 \pm 0.101$ | $0.057 \pm 0.113$ | 0.0001 |
| PA pressure |  |  |  |  |
| Systolic(mmHg) | $18.6 \pm 2.8$ | $61.3 \pm 17.0$ | $23.4 \pm 8.6$ | <0.0001 |
| Diastolic (mmHg) | $9.2 \pm 2.0$ | $32.3 \pm 9.8$ | $8.2 \pm 3.7$ | <0.0001 |
| Mean (mmHg) | $13.2 \pm 2.0$ | $44.1 \pm 11.9$ | $14.4 \pm 4.5$ | <0.0001 |
| RAP (mmHg) | $5.6 \pm 1.4$ | $9.0 \pm 2.4$ | $7.8 \pm 2.2$ | 0.0002 |
| LV pressure |  |  |  |  |
| LVSP(mmHg) | 82.6 ± 8.3 | 82.3 ± 10.4 | 83.6 ± 10.6 | 0.7797 |
| LVEDP(mmHg) | 10.5 ± 2.6 | 10.7 ± 3.5 | 10.7 ± 2.4 | 0.9597 |
| Ao pressure |  |  |  |  |
| Systolic(mmHg) | 82.6 ± 7.6 | 77.2 ± 12.0 | 83.4 ± 11.1 | 0.4471 |
| Diastolic(mmHg) | 52.7 ± 7.2 | 49.6 ± 6.3 | 51.4 ± 9.9 | 0.5949 |
| Mean(mmHg) | 65.6 ± 6.3 | 62.0 ± 9.4 | 65.1 ± 10.7 | 0.6673 |

BSA, body surface area; HR, heart rate; RV, right ventricle; RVSP, right ventricular systolic pressure; Pmin, right ventricular minimum pressure; RVEDP, right ventricular end-diastolic pressure; dP/dt\_max, right ventricular maximum pressure derivative; dP/dt\_min, right ventricular minimum pressure derivative; AugPr, Augmentation Pressure; PA, pulmonary artery; RAP, right atrial pressure; LV, left ventricle; LVSP, left ventricular systolic pressure; LVEDP, left ventricular end-diastolic pressure; Ao, aorta

No significant differences in age, height, body weight, body surface area, or heart rate were found among the groups. Parameters reflecting RV function, LV function, and pulmonary circulatory status demonstrated findings consistent with the pathophysiological characteristics of each disease group, as shown in Table 1.

### PPP and AugPr in each study group

Before evaluating AugPr and the AugPr index using PPP in the rTOF cohort, we first examined PPP findings and PA reflection in the PAH group—where PA wave reflection has previously been reported—and in the control group. Representative RV pressure waveforms and PPP plots for the control and PAH groups are shown in Fig 1. No AugPr was observed in the control group. By contrast, in a representative recording of patients in the PAH group, AugPr was 20 mmHg and the AugPr index was 0.32. AugPr and the AugPr index were easily quantifiable and visually identifiable on PPP analysis.

### PPP and AugPr in patients with rTOF

Next, we evaluated PPP and AugPr in the rTOF cohort. Patient characteristics according to surgical procedure are shown in Table 2. No significant differences in age, height, body weight, body surface area, or heart rate were observed among the PVS, TAP, and Rastelli groups. RVSP, dP/dt_max, and dP/dt_min were significantly higher in the Rastelli group than the PVS and TAP groups (p < 0.004, 0.0003, and 0.0002, respectively). No significant differences in Pmin or Tau were observed among the three groups. PA pressure, right atrial pressure, and systemic arterial pressure did not differ significantly among the groups.

**Table 2.** The clinical and hemodynamic characteristics of r-TOF subgroups.

| Parameter | PVS | TAP | Rastelli | p |
| --- | --- | --- | --- | --- |
| n | 5 | 34 | 48 |  |
| Age (years) | 10.8 ± 9.5 | 16.1 ± 12.5 | 12.0 ± 7.9 | 0.2758 |
| Height (cm) | 125.9 ± 41.8 | 138.5 ± 40.1 | 131.2 ± 31.1 | 0.6525 |
| Weight (kg) | 32.8 ± 22.2 | 41.8 ± 23.1 | 35.0 ± 18.6 | 0.3869 |
| BSA (m<sup>2</sup>) | 1.05 ± 0.56 | 1.24 ± 0.55 | 1.11 ± 0.44 | 0.5232 |
| HR (bpm) | 93.5 ± 20.4 | 85.7 ± 28.9 | 86.7 ± 14.5 | 0.7629 |
| RV pressure |  |  |  |  |
| RVSP (mmHg) | 35.0 ± 7.4 | 35.4 ± 10.2 | 55.4 ± 23.3<sup>1)</sup> | 0.0004 |
| Pmin (mmHg) | 2.0 ± 2.1 | 2.6 ± 1.7 | 2.8 ± 2.2 | 0.3446 |
| RVEDP (mmHg) | 8.0 ± 2.3 | 8.6 ± 2.2 | 9.0 ± 2.5 | 0.5516 |
| dP/dt max<br>(mmHg/s) | 284 ± 36.6 | 315.2 ± 146.1 | 424.6 ± 175.3<sup>2)</sup> | 0.0003 |
| dP/dt min<br>(mmHg/s) | -319.2 ± 71.6 | -245.1 ± 173.1 | -469.7 ± 207.2<sup>3)</sup> | 0.0002 |
| Tau (ms) | 46.0 ± 17.3 | 47.7 ± 14.4 | 48.4 ± 12.5 | 0.9298 |
| AugPr (mmHg) | 0 ± 0 | 0.6 ± 1.7 | 5.7 ± 10.8 | 0.4555 |
| AugPr index | 0 ± 0 | 1.2 ± 3.7 | 9.2 ± 15.8 | 0.5578 |
| PA pressure |  |  |  |  |
| Systolic (mmHg) | 18.8 ± 3.0 | 23.1 ± 7.6 | 27.0 ± 11.0<sup>4)</sup> | 0.0031 |
| Diastolic (mmHg) | 7.8 ± 2.7 | 6.9 ± 3.0 | 8.5 ± 3.9 | 0.1659 |
| Mean (mmHg) | 13.2 ± 2.3 | 13.6 ± 2.7 | 12.2 ± 6.0 | 0.4047 |
| RVOTO (mmHg) | 16.2 ± 8.8 | 12.4 ± 8.6 | 28.3 ± 22.8<sup>5)</sup> | 0.0020 |
| RAP (mmHg) | 7.2 ± 1.6 | 7.4 ± 1.7 | 8.7 ± 2.2<sup>6)</sup> | 0.0291 |
| LVSP (mmHg) | 84.6 ± 12.3 | 81.8 ± 13.2 | 82.9 ± 10.7 | 0.8725 |
| LVEDP (mmHg) | 10.4 ± 3.5 | 11.4 ± 2.2 | 11.3 ± 2.5 | 0.8336 |
| Ao pressure |  |  |  |  |
| Systolic (mmHg) | 83.0 ± 13.5 | 83.0 ± 13.8 | 82.9 ± 12.0 | 0.9994 |
| Diastolic (mmHg) | 49.4 ± 9.6 | 53.5 ± 13.6 | 49.5 ± 8.7 | 0.3668 |
| Mean (mmHg) | 64.2 ± 15.4 | 67.3 ± 15.2 | 63.4 ± 9.5 | 0.4653 |
| LV and RV<br>volume |  |  |  |  |
| LVEDVI (mL/m<sup>2</sup>) | 78.5 ± 17.6 | 82.4 ± 20.1 | 89.3 ± 14.1 | 0.1750 |
| %LVEDV (%) | 88.9 ± 7.5 | 100.1 ± 24.6 | 111.3 ± 27.4<sup>7)</sup> | 0.0011 |
| LVEF (%) | 61.7 ± 15.9 | 58.3 ± 9.2 | 58.9 ± 10.4 | 0.8862 |
| RVEDVI (mL/m<sup>2</sup>) | 95.7 ± 16.9 | 112.6 ± 39.2 | 105.9 ± 35.5 | 0.2894 |
| %RVEDV (%) | 118.1 ± 17.0 | 124.9 ± 44.1 | 121.8 ± 42.1 | 0.8252 |
| RVEF (%) | 55.0 ± 8.7 | 51.7 ± 9.6 | 46.8 ± 11.8 | 0.0915 |
| PR grade | 1.0 ± 1.0 | 1.8 ± 1.6 | 1.9 ± 1.5 | 0.2437 |

BSA, body surface area; HR, heart rate; RV, right ventricle; RVSP, right ventricular systolic pressure; Pmin, right ventricular minimum pressure; RVEDP, right ventricular end-diastolic pressure; dP/dt\_max, right ventricular maximum pressure derivative; dP/dt\_min, right ventricular minimum pressure derivative; AugPr, Augmentation Pressure; PA, pulmonary artery; RVOTO, right ventricular outflow tract obstruction; RAP, right atrial pressure; LV, left ventricle; LVSP, left ventricular systolic pressure; LVEDP, left ventricular end-diastolic pressure; Ao, aorta; LVEDVI, left ventricular end-diastolic volume index; %LVEDV, % normal of left ventricular end-diastolic volume; LVEF, left ventricular ejection fraction; RVEDVI, right ventricular end-diastolic volume index; %RVEDV, % normal of right ventricular end-diastolic volume; RVEF, right ventricular ejection fraction, PR, pulmonary regurgitation; PR grade, 0=none, 1= trivial, 2= mild, 3= moderate, 4= severe
1) 0.0018 vs. PVS and <0.0001 vs. TAP; 2) 0.0002 vs. PVS and 0.0089 vs. TAP; 3) 0.0128 vs. PVS and <0.0001 vs. TAP; 4) 0.0024 vs. PVS; 5) 0.0208 vs. PVS and <0.0001 vs. TAP 6) 0.0103 vs. TAP; 7) 0.0010 vs. PVS

PPP, AugPr, and the AugPr index in the rTOF group are shown in Fig 2. Among the patients in the rTOF group, both cases with and without detectable PA wave reflection were observed. No AugPr was observed in the PVS group (Fig 2B). In the representative recording of the Rastelli group case shown in Fig 2D, AugPr was 12 mmHg and the AugPr index was 0.25.

**Fig 2.**
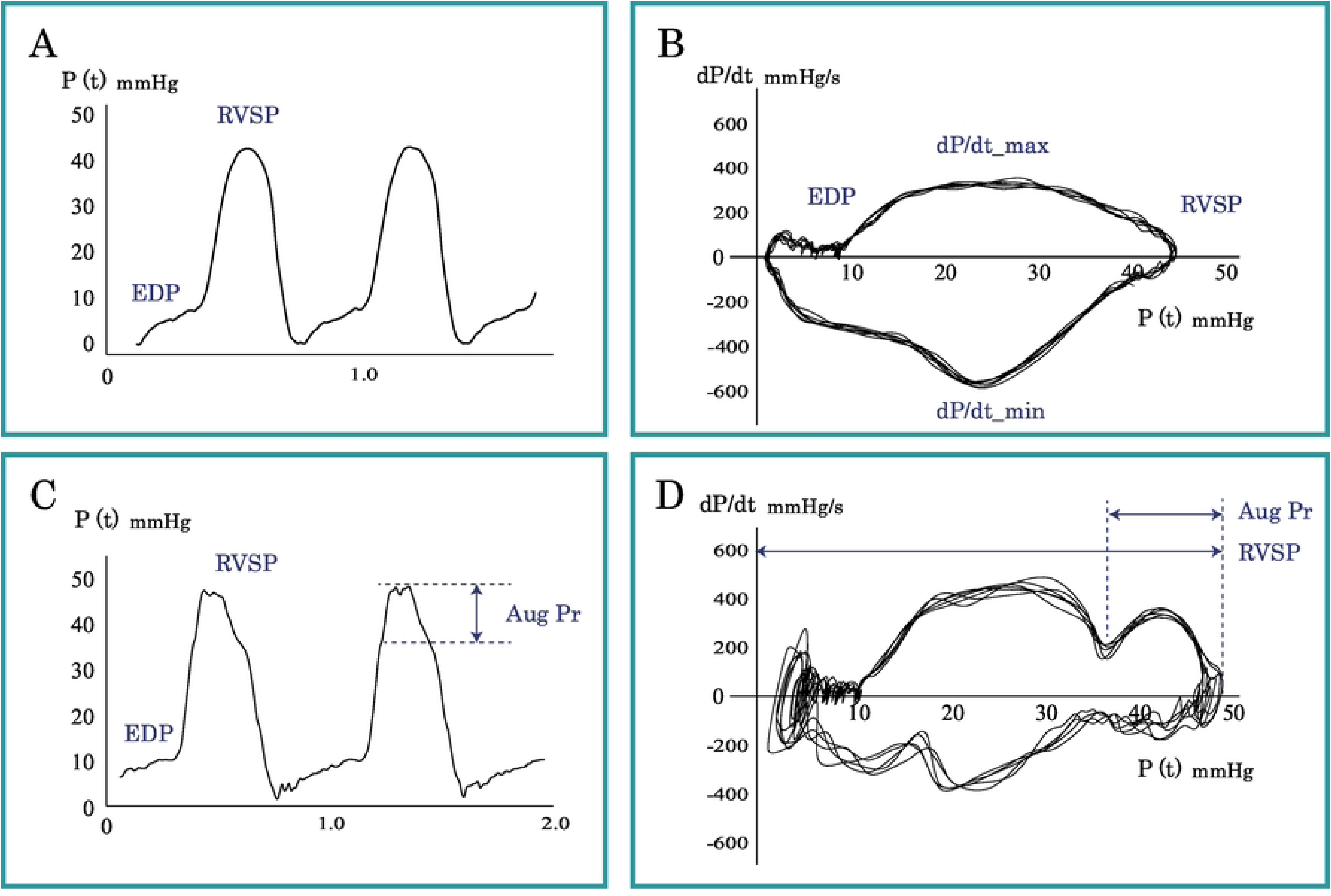
Right ventricular (RV) pressure waveform and Pressure phase plane (PPP) of RV pressure in the repaired tetralogy of Fallot (rTOF) group. A. Representative recording of RV pressure waveform from a patient in the pulmonary valve–sparing repair (PVS) subgroup of the repaired tetralogy of Fallot (rTOF) cohort. B. Pressure phase plane (PPP) of the right ventricular (RV) pressure from the same patient with PVS shown in A. No late systolic pressure augmentation is observed. C. Representative recording of an RV pressure waveform from a patient in the Rastelli subgroup of the rTOF cohort. D. PPP of the RV pressure from the same patient in the Rastelli group shown in C. In this case, AugPr is 12 mmHg and the AugPr index is 0.25. RVSP, right ventricular systolic pressure; EDP, end-diastolic pressure; AugPr, augmentation pressure.

### Evaluation of pulmonary wave reflection, AugPr, and the AugPr index among the three groups

The prevalence of AugPr in the three study groups is shown in Fig 3A. The PAH group demonstrated a significantly higher prevalence of AugPr than the other two groups, followed by the rTOF group, whereas no cases were observed in the control group. A significant difference in prevalence was confirmed among the three groups (p < 0.0001). AugPr and the AugPr index for the three groups are shown in Fig 3B and 3C. The PAH group demonstrated significantly higher AugPr and AugPr index values compared with the control and rTOF groups (both p < 0.0001).

**Fig 3.**
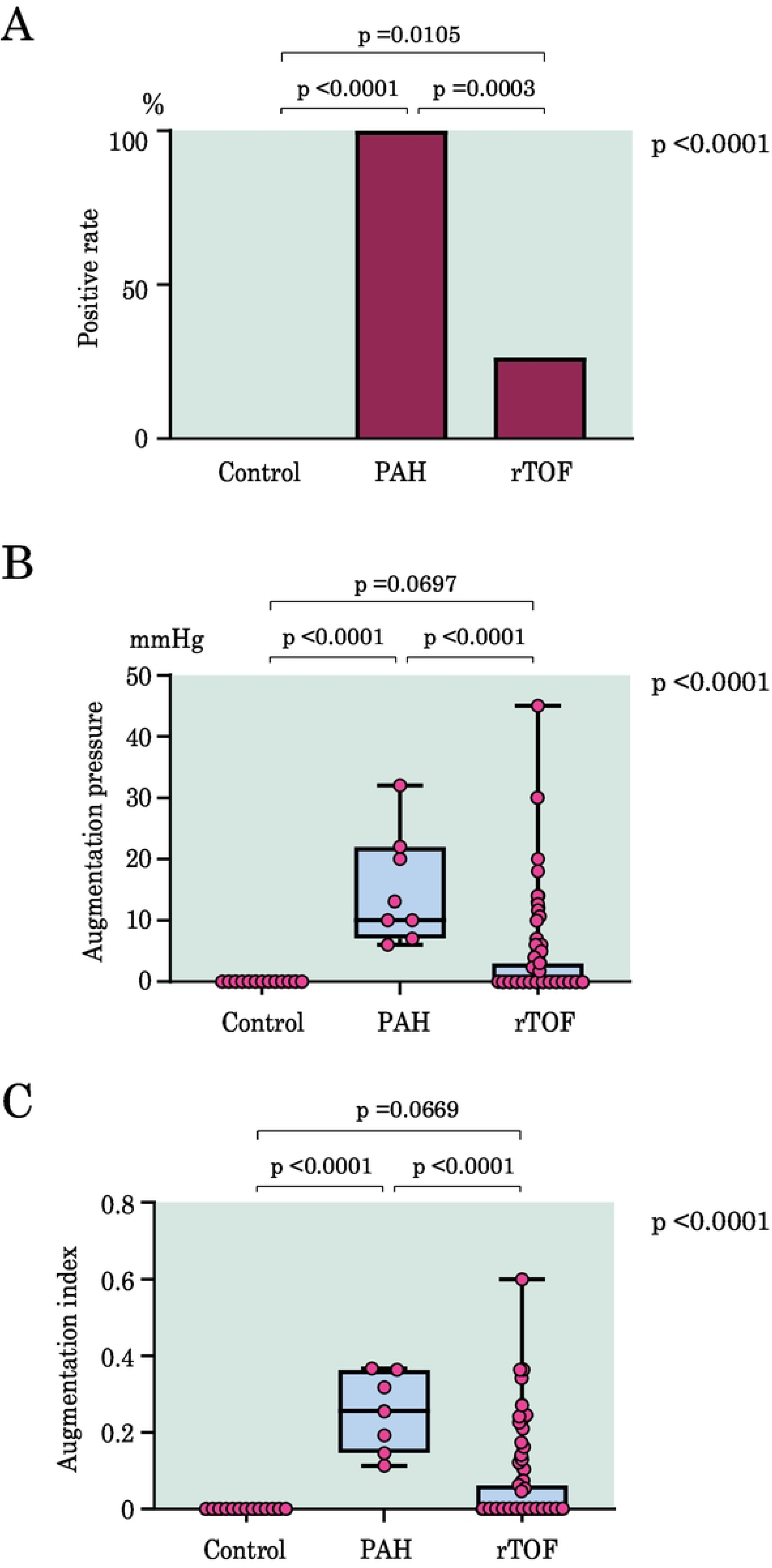
Comparison of AugPr Prevalence and Indices Among Control, PAH, and rTOF Groups. A. Prevalence of AugPr positivity in the control, PAH, and rTOF groups. B. AugPr values in the three groups. Red circles indicate individual values; boxes represent the interquartile range (IQR); the central line indicates the median; whiskers extend from minimum to maximum. C. AugPr index values in the three groups. Red circles indicate individual values; boxes represent the IQR; the central line indicates the median; whiskers extend from minimum to maximum.

### Pulmonary wave reflection, AugPr, and the AugPr index according to surgical procedure in the rTOF group

The 87 patients in the rTOF group were classified into three subgroups according to surgical technique: PVS group (n = 5), TAP group (n = 34), and Rastelli group (n = 48). The Rastelli group demonstrated a significantly higher prevalence of AugPr (Fig 4A) (p = 0.0027). Furthermore, AugPr and the AugPr index were significantly higher in the Rastelli group than in the PVS and TAP groups (Fig 4B and 4C) (p = 0.0447 and 0.0433, respectively).

**Fig 4.**
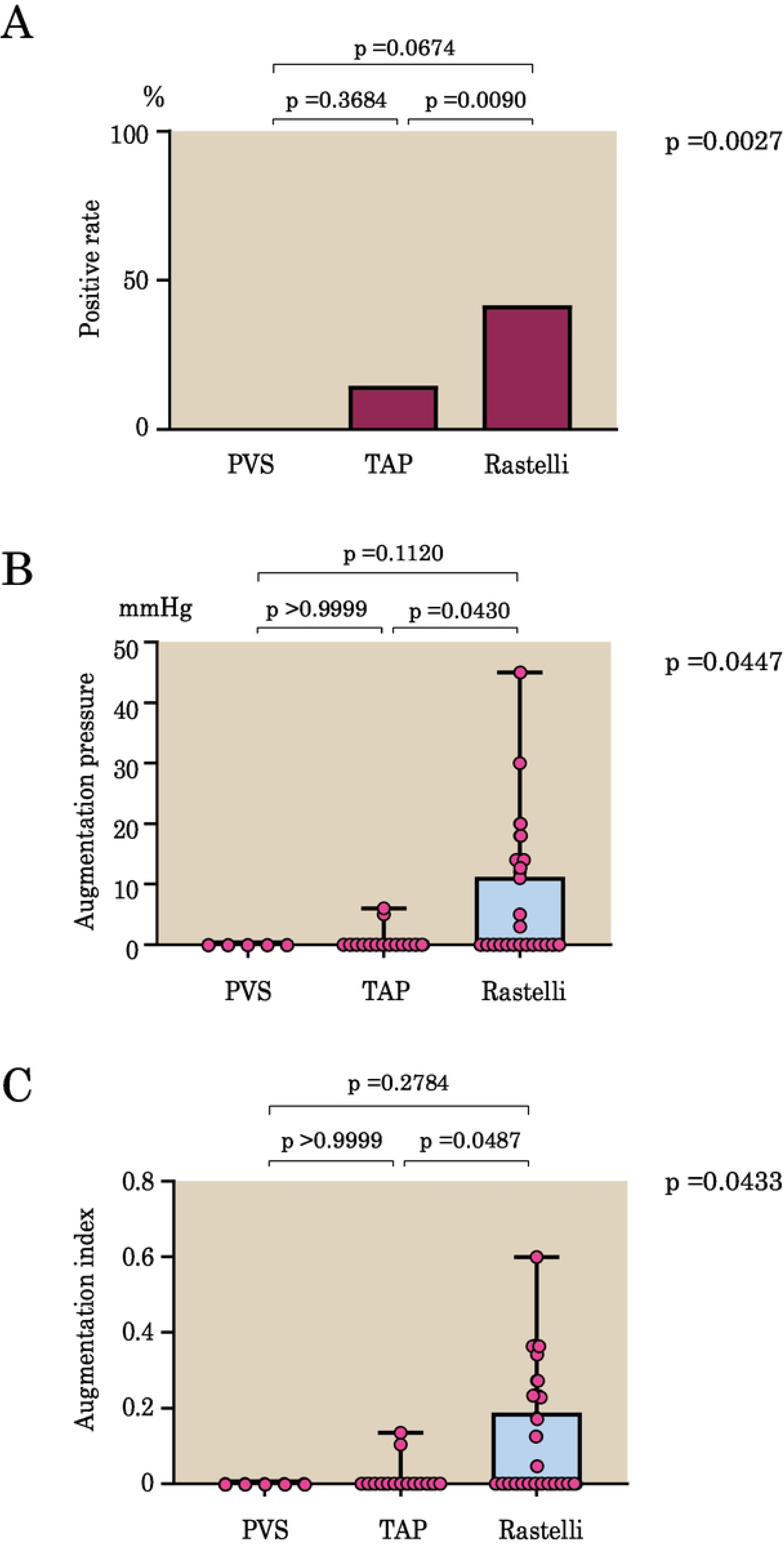
Comparison of AugPr Measures Among PVS, TAP, and Rastelli Subgroups. A. Prevalence of AugPr positivity in the PVS, TAP, and Rastelli subgroups. B. AugPr values in the three surgical subgroups. Red circles indicate individual values; boxes represent the interquartile range (IQR); the central line indicates the median; whiskers extend from minimum to maximum. C. AugPr index values in the three surgical subgroups. Red circles indicate individual values; boxes represent the IQR; the central line indicates the median; whiskers extend from minimum to maximum.

### Relationship between AugPr, the AugPr index, and hemodynamic parameters in the rTOF group

Fig 5 shows the relationships between AugPr, the AugPr index, and hemodynamic parameters in the rTOF cohort. The analyzed hemodynamic parameters are listed in Table 2 and include both right- and left-sided indices. Among these variables, RVSP, RVOTO, and dP/dt_max showed significant correlations with both AugPr and the AugPr index. PR grade was significantly correlated with AugPr, but not with the AugPr index.

**Fig 5.**
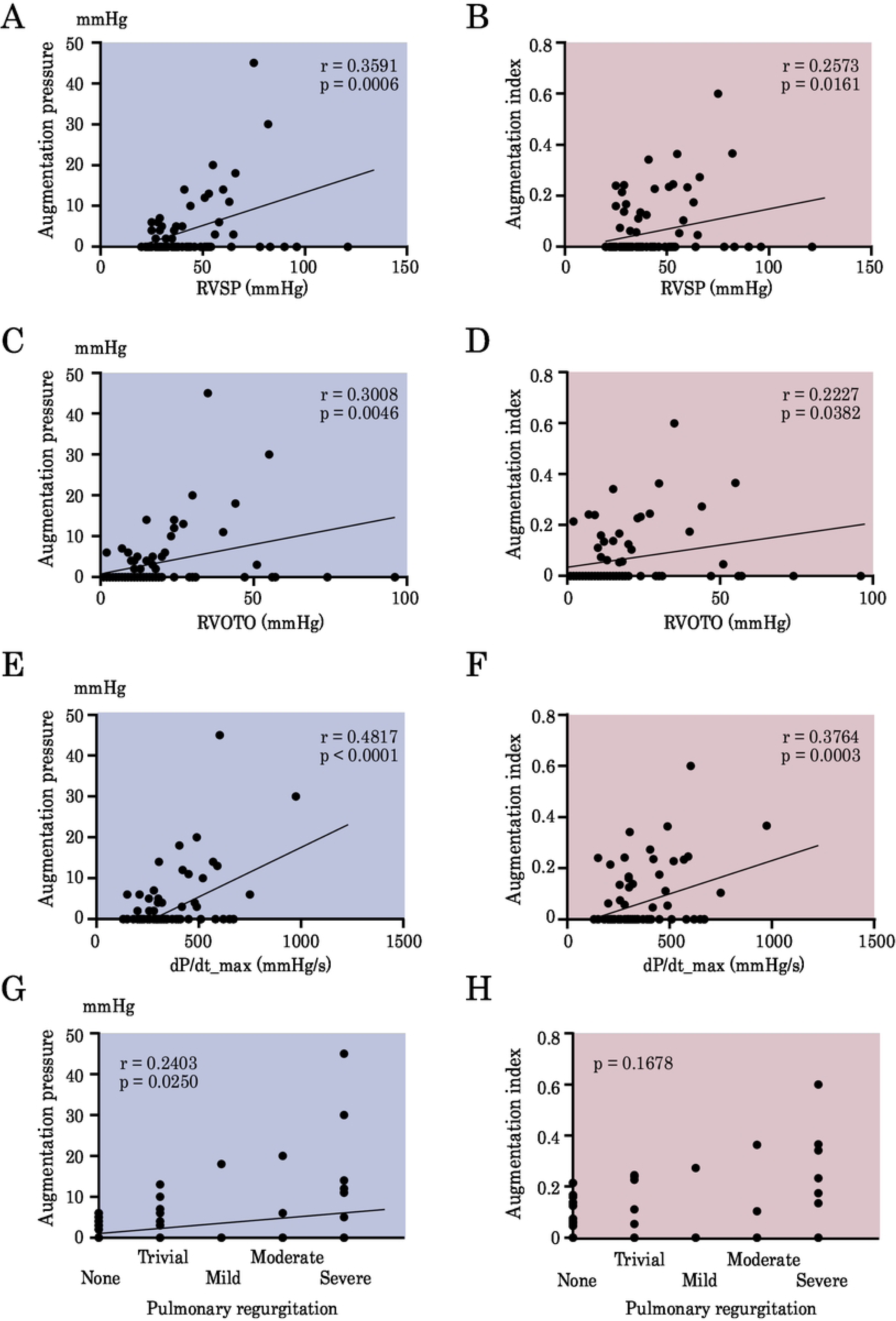
Relationships between AugPr and the AugPr index and hemodynamic parameters. Significant correlations were observed between AugPr/AugPr index and RVSP (A, B), RVOTO (C, D), and dP/dt_max (E, F). A significant correlation was observed between PR grade and AugPr (G), whereas no significant correlation was found between PR grade and the AugPr index (H).

### Reproducibility

To assess the reproducibility of AugPr and the AugPr index, intra- and interobserver variabilities in the measurements were confirmed in 10 randomly selected participants (rTOF [n = 7] and PAH [n – 3]) by means of ICCs and Bland–Altman analysis. These 10 cases were selected from those with positive AugPr results. The ICCs of AugPr were 0.991 and 0.995 for intra- and interobserver variability, respectively. The ICCs of the AugPr index for intra- and interobserver reproducibility were 0.943 and 0.978, respectively. Bland–Altman analysis also showed minimal bias and substantial agreement for reproducibility (Fig 6). AugPr and AugPr index measurements proved to be highly reproducible.

**Fig 6.**
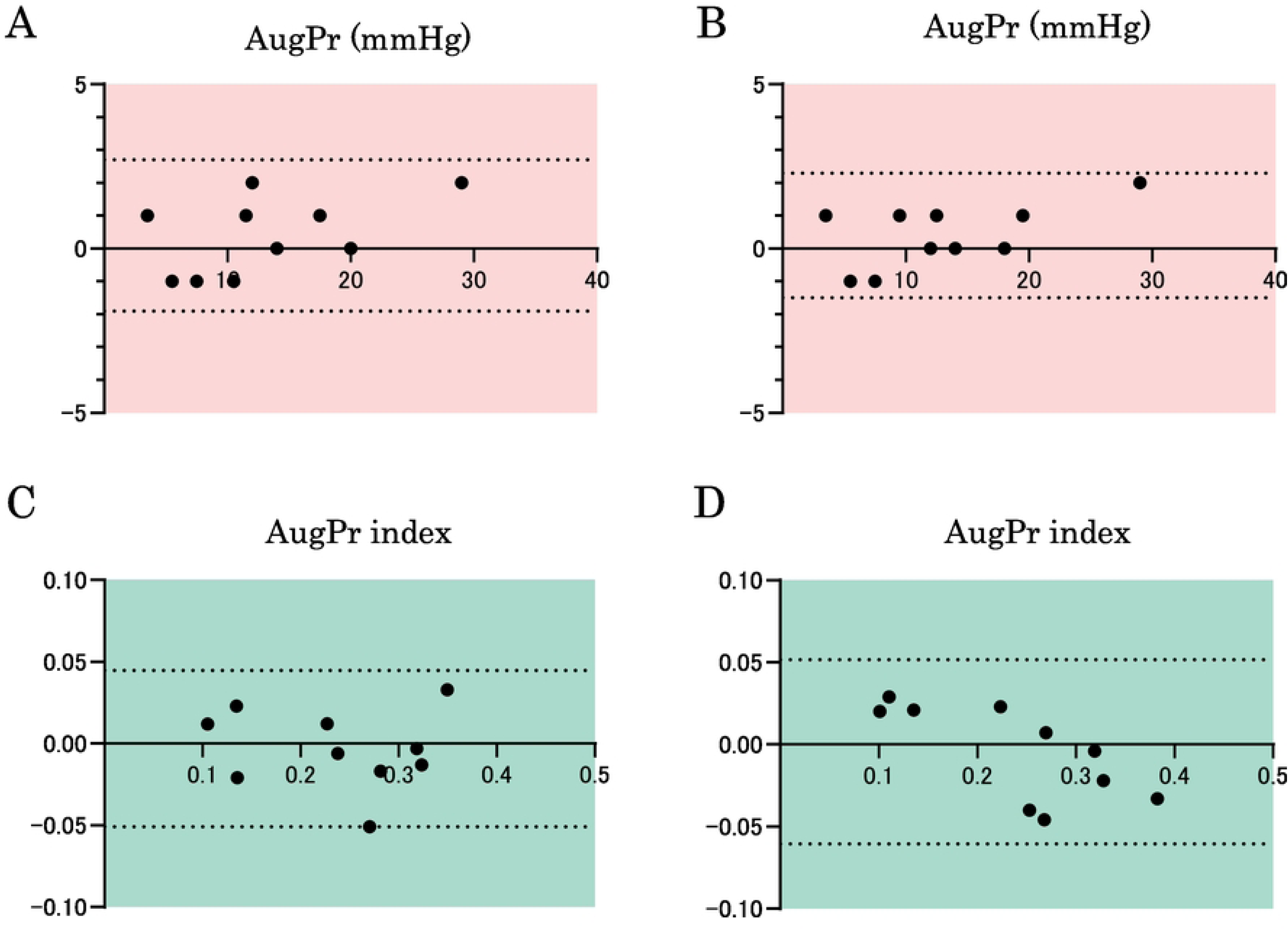
Bland–Altman plots for the parameters of AugPr and AugPr index. Intraobserver (A and C) and interobserver (B and D) variabilities are shown. The dotted lines show the bias ± 1.96 SD (95% limit of agreement).

## Discussion

In the present study, we demonstrated that the magnitude of PA wave reflection can be assessed from the configuration of the PPP loop using the RV pressure waveform. Furthermore, we confirmed that this method can quantitatively assess PA wave reflection, which was previously difficult to evaluate clinically in patients with rTOF because of PR. Among the different surgical procedures for rTOF, wave reflection was most prominent in the Rastelli group.

Previous investigations of PA wave reflection in the right heart have primarily been conducted in patients with PAH, where its utility for prognostic assessment has been reported [5,6,17,18]. However, compared with studies involving systemic circulation, reports focusing on the right heart are limited, and to our knowledge, no studies have been reported in patients with congenital heart disease. In patients with PAH, late systolic peaking of the RV pressure waveform is not uncommon [17,18]. This phenomenon is considered to result from wave reflection rather than dynamic RVOTO and has been reported to resemble late systolic peaking in the left heart. The configuration of the PPP loop concisely reflects how the RV responds to wave reflection and increased afterload. In PAH, increased PA stiffness and wave reflection result in a reflected wave returning to the RV during late systole, producing a secondary systolic peak [5,6,17,18]. Some studies have shown that late systolic peaking is more pronounced in the RV pressure waveform than in the PA pressure waveform in patients with PAH [18,19]. Although this differs from the characteristics observed in systemic circulation, the precise mechanism remains unclear. These right-sided hemodynamic properties support the clinical relevance of evaluating PA wave reflection using PPP analysis of the RV pressure waveform, as performed in the present study.

When wave reflection is assessed using the aortic pressure waveform, some reports define AugPr at the point where the fourth derivative of the pressure waveform becomes zero (corresponding to an inflection point in the third derivative). Although the systolic inflection point identified on PPP analysis is not strictly identical to this definition, the difference is unlikely to be clinically significant. Kelly et al. [20] proposed the use of the fourth derivative waveform to enable objective and automated measurement of the AIx. They defined the timing of early systolic inflection as the second zero-crossing point where the fourth derivative waveform crosses from positive to negative. Takazawa et al. [21] reported the use of the third zero-crossing of the fourth derivative waveform for this purpose. Thus, even in the definition of AugPr in the left heart system, there is no single strict definition.

In the rTOF cohort, RVSP, dP/dt_max, RVOTO, and PR grade were significantly correlated with the magnitude of PA wave reflection. It is possible that stronger wave reflection increased RV afterload, thereby resulting in higher RVSP and dP/dt values. Alternatively, because these hemodynamic characteristics were more pronounced in the Rastelli group, the observed correlations may reflect the specific pathophysiology of this subgroup. Among patients in rTOF group, the Rastelli group demonstrated both the highest prevalence and the greatest magnitude of wave reflection. Pulmonary wave reflection in patients with rTOF may be influenced by factors such as the angle, curvature, and stiffness of the prosthetic conduit, as well as the branching geometry and impedance characteristics of the right and left PA.

### Limitations

This study has some limitations. First, the sample size was relatively small. A larger cohort is required to determine the prevalence and magnitude of pulmonary wave reflection more accurately in each disease group. In particular, patients in the rTOF group exhibited substantial heterogeneity in terms of the clinical course, treatment strategies, and RV and PA morphology. Although we classified surgical procedures into three categories, each category encompasses diverse operative techniques. Larger studies are needed to clarify which specific pathophysiological factors influence pulmonary wave reflection most strongly in patients with rTOF. Second, there are limitations regarding the diagnostic and evaluative methodology of PPP analysis. If the reflected wave returns very early or very late during systole, it may not appear as the late systolic pressure augmentation pattern described in this study, and may not be adequately detected by PPP analysis. In such cases, a clear inflection point in systolic dP/dt may not be observed. Conversely, RV pressure waveforms influenced by RVOT stenosis, pulmonary valve motion, or tricuspid valve abnormalities may potentially mimic patterns interpreted as wave reflection on PPP analysis. It is necessary to recognize that there are certain limitations to evaluating AugPr using PPP in this manner.

## Conclusion

The findings of the present study indicate that RV PPP analysis can be used to evaluate PA wave reflection in patients with rTOF. This method provides clear visualization and represents a logical and clinically meaningful approach to assessment. Among the surgical procedures, PA wave reflection was the most pronounced in the Rastelli group. Furthermore, significant associations were found among RVSP, dP/dt_max, RVOTO, and PR grade.

## Data Availability

The minimal anonymized dataset underlying the findings of this study, together with the analysis code used to generate the results, is available in Figshare at https:/doi.org/10.6084/m9.figshare.32155395. All data have been fully anonymized and de-identified in accordance with institutional and ethical guidelines.

https://doi.org/10.6084/9.figshare.32155395

## Data Availability

All relevant data are available from the figshare repository (doi: xxx).

